# Clinical Characteristics and In-Hospital Outcomes of Acute Heart Failure Patients In Ethiopia: A Retrospective Cohort Study

**DOI:** 10.1101/2025.05.29.25328611

**Authors:** Wubshet Abraham Alemu, Duresa Mieso Elemo, Tamiru Adugna Dadi, Koricho Simie Tolla, Ayalneh Demissie Ashenafi

## Abstract

**Background:** Acute heart failure (AHF) is a critical cardiovascular emergency with substantial mortality in low-resource settings. This study examines the clinical characteristics and outcomes of acute heart failure patients in Ethiopia. The burden in low-resource settings like Ethiopia remains under-characterized.

**Methods:** An institution-based retrospective cohort study was conducted at Asella Referral and Teaching Hospital, Ethiopia from September 2022 to August 2023. Patients admitted with acute heart failure during study period were included. Data were extracted from medical charts and analyzed with SPSS version 28. Descriptive statistics summarized baseline characteristics. Kaplan–Meier survival curves and long-rank tests assessed survival distributions. Cox proportional hazards models identified mortality predictors.

**Results:** Among 231 patients, 52.4% were female. The median age was 56 years (IQR 45-68). The leading precipitating factor was pneumonia (24.68%), and ischemic heart disease (36.4%) was the most common underlying condition. In-hospital mortality was 13.4%. Pneumonia (HR 3.07; 95% CI: 1.40-6.74), acute kidney injury (HR 2.98; 95% CI: 1.31-6.75), lack of health insurance (HR 3.46; 95% CI: 1.51-7.95), and age >65 years (HR 2.81; 95% CI: 1.18-6.72) were independently associated with mortality

**Conclusion:** The high in-hospital mortality among acute heart failure was significantly associated with pneumonia, acute kidney injury, lack of health insurance, and advanced age. Targeted interventions addressing these factors may improve outcomes.

## 1. Introduction

Heart failure is a clinical syndrome where the heart is unable to pump enough blood to meet the body’s demand, resulting in observable symptoms and signs. Acute heart failure refers to the sudden or rapid onset of symptoms and signs of heart failure that require urgent medical attention (1,2).

Commonly presented symptoms include dyspnea during exercise or at rest, orthopnea, fatigue, and reduced exercise tolerance. Clinical signs of congestion, such as peripheral edema, jugular vein distension, and S3 gallop, often accompany symptoms. AHF is classified into two groups according to the presence or absence of previous HF: worsening (decompensated) preexisting stable HF suddenly or progressively described as decompensated AHF and new (de novo) HF, where there is no known previous HF (1–3).

Based on the presence of signs of congestion and/or peripheral hypoperfusion, there are four major clinical presentations of acute heart failure, with possible overlaps between them: acutely decompensated heart failure, acute pulmonary edema, isolated right ventricular failure, and cardiogenic shock (1).

The diagnosis requires the presence of symptoms and/or signs of AHF and supporting evidence from investigations such as signs of pulmonary edema and cardiomegaly on chest X-ray, echocardiogram (ECHO) findings, and elevated cardiac biomarkers like BNP (≥100 pg/mL), NT-proBNP (≥300 pg/mL), MR-proANP (≥120 pg/mL), and ECG (1).

The goal of treatment should be to reverse acute hemodynamic abnormalities, quickly relieve symptoms, and initiate treatments that will decrease disease progression and improve survival. The management can be simplified and improved by assessing the most likely hemodynamic profile based on clinical signs and symptoms (1,4).

Patients with heart failure who are admitted with evidence of significant fluid overload should be treated with intravenous loop diuretics to improve symptoms and reduce morbidity. In addition, vasodilation therapy may be considered an adjuvant to diuretic therapy for the relief of dyspnea. In patients with cardiogenic shock, intravenous inotropic support should be used to maintain systemic perfusion and preserve end-organ perfusion. It is also important to monitor the patient’s oxygen levels and provide oxygen for patients with hypoxia. In addition, hospitalization is a critical opportunity to continue, initiate, and further optimize GDMT (4). Research conducted in developing nations indicates that heart failure (HF) is usually precipitated by avoidable factors such as infections, uncontrolled hypertension, and arrhythmias, as well as anemia. Moreover, a considerable proportion of acute HF patients are relatively young, with a median age below 55 years, experiencing unsatisfactory in-hospital prognoses. This circumstance may be attributed to a combination of challenges, including restricted access to healthcare services, insufficient insurance coverage, and inadequacies in healthcare infrastructure and quality (5,6).

In sub-Saharan Africa, patients tend to present at a younger age and with severe symptoms. Common precipitating factors include infections, arrhythmia, and poor medication adherence. There are limited data on predictors of acute heart failure mortality in Ethiopian tertiary centers. This study investigates the characteristics and outcomes of acute heart failure outcomes in a tertiary hospital setting in Ethiopia. The goal is to focus attention on the major factors that were reasons for admission as it will hep identify patients with possible predictors of poor outcome.

## 2. Methods and Materials

### 2.1 Study Design and Setting

A retrospective cohort study was conducted at Asella Referral and Teaching Hospital (ARTH), affiliated with Arsi University. The hospital serves over 3.5 million people in southeast Ethiopia. Asella town is located 175 km southeast of the capital, Addis Ababa. The estimated population of the town at mid-2022 was 139,537, of whom 69,459 were male and 70,078 were female (45). The hospital has a capacity of over 200 beds, with the medical ward comprising approximately 40–50 of these, ICU (6 beds), and HDU (4 beds). The medical ward provides inpatient care for a wide range of internal medicine cases, including infectious diseases (such as TB and HIV/AIDS), chronic diseases (such as diabetes, hypertension, and heart failure), and acute medical emergencies.

The hospital is staffed with general practitioners, cardiologists, internal medicine specialists, residents, interns, nurses, and medical students. It is equipped with diagnostic facilities, including laboratory services, radiology, and pharmacy services, although resources may sometimes be limited.

Data were collected between November 1, 2023, and January 31, 2024, covering patients admitted from September 2022 to August 2023.

Authors had no access to information that could identify individual participants during or after data collection.

### 2.2 Source and study population

The source population was patients admitted to the medical ward, ICU, and HDU of ARTH. The study population was patients admitted to the medical ward, ICU, and HDU of ARTH with a diagnosis of AHF during the study period.

### 2.3 Eligibility criteria

#### 2.3.1 Inclusion criteria

Patients Diagnosed with Acute Heart Failure (AHF).

Patients admitted for duration of exceeding 24 hours.

Patients aged 18 years or older

#### 2.3.2 Exclusion criteria

Patients with an unknown outcome.

Patients with advanced malignancy.

Patients with stage 4 HIV infection.

### 2.4 Sample size determination

All eligible patients admitted within the one-year period were included.

### 2.5 Data collection

Trained nurses extracted data using a structured format based on national and international guidelines (1,4,48). Extracted data included demographics, clinical features, lab results, imaging, precipitating factors, underlying conditions, treatments, and outcomes.

### 2.6 Variables of the study

#### 2.6.1 Dependent variable

The primary outcome of our research is in-hospital mortality. Secondary outcomes include patients leaving against medical advice and improvements in health status at discharge, measured by improved NYHA functional class and resolution of edema. The study endpoint is the duration in days from admission to events such as death, discharge with the same condition, improvement, or leaving against medical advice.

#### 2.6.2 Independent variables

- **Socio-demographic:** age, gender, insurance coverage,
- **Life-styles variables**: smoking, alcohol drinking.
- **Vital signs** (PR, BP, PR, SPo2, Temperature, Urine output),
- **Framingham major and minor criteria**,
- **Heart failure syndrome** (De novo, Acute decompensated heart failure)
- **Hemodynamic profile** (CS, PE, ADHF, RHF)
- **Number of readmission**
- **Imaging study** (ECHO, ECG, CXR,)
- **Laboratory values** (serum Na, serum K, creatinine, BUN, Hemoglobin, Troponin),
- **Precipitating factors:** infection, noncompliance with medication, noncompliance with salt restriction, anemia, ACS, uncontrolled HTN, arrhythmia, pulmonary embolism, pregnancy, worsening of renal function.
- **Underlying disease:** IHD, VHD, cardiomyopathy, HHD, Cor-pulmonale, degenerative valvular heart disease, congenital heart disease.
- **Co-morbid variables:** CKD, DM, HIV/AIDS, HTN, COPD, Dyslipidemia, Cancer, Arrhythmia.
- **Drug use assessment:** the medication the patient is on before admission, during admission, and at discharge (beta blocker, ACEI, ARBS, aspirin, atorvastatin, mineralocorticoid receptor antagonist, adenosine, amiodarone, calcium channel blocker, digoxin, diuretics, dopamine, labetalol, norepinephrine, or vasodilator) and adverse drug reactions/side effects.

### 2.7 Operational definitions

**Acute heart failure:** signs and symptoms of new onset of HF and/or decompensation or worsening of chronic stable HF.

**Improved:** is defined as hemodynamically stable, decongested, started on oral evidence-based medication, and tolerated for 24 hr.

**In hospital mortality:** is defined as death occurring during hospitalization for acute heart failure

**Heavy drinking:** for men, consuming more than 15 drinks per week; for women, more than 8 drinks per weak (47).

**Moderate drinking:** 2 drinks or less in a day for men, 1 drink or less in a day for women (47).

**Poor outcome:** the attainment of one of the following end results: death and self-discharge against medical advice with no improvement.

**Readmission:** is when the same patient was admitted within the study period.

**Smoker:** A patient who has smoked 100 cigarettes or more in his or her lifetime and who currently smokes (46).

**The same:** if there is no significant change in hemodynamic status (requiring IV diuretics, vasopressor, inotropes, HR>100 beat/min or patient in shock) or congested during discharge.

**Treatment outcome:** the attainment of a specified end result measured using parameters such as improved and/or died.

### 2.8 Data collection procedures

The data extraction checklist for this study was meticulously designed in accordance with established guidelines, including the Ethiopian National Guideline on major non-communicable diseases (2016), the European Society of Cardiology (ESC) guidelines from 2021, and the ACC/AHA guidelines and definitions (1,4,48). This comprehensive checklist includes a range of crucial elements such as socio-demographic details, clinical characteristics, precipitating factors, underlying diseases, co-morbidities, and imaging studies, including chest X-ray, electrocardiogram, and echocardiography. Additionally, the checklist captures essential information on the treatment administered and the duration of the hospital stay. Furthermore, the data extraction process involved the systematic collection of vital signs and laboratory values, including serum sodium, potassium, hemoglobin, serum creatinine, blood urea nitrogen (BUN), troponin, and the estimated glomerular filtration rate (GFR) based on the CKD-EPI-derived formula. We evaluate the outcome from the last admission. Data was collected using a pretested format by two trained nurses. Relevant clinical information and data were obtained from patient charts.

### 2.9 Data quality assurance

One day of training was given for data collectors before entering into the data collection process on the objective and relevance of the study, how to gather the appropriate information, procedures of data collection techniques, and the whole contents of the checklist. The data collection process and completeness were closely supervised. A pre-test was done to assure clarity, avoidance of ambiguity, comprehensiveness, and content uniformity.

### 2.10 Data processing and analysis

The collected data, after being manually checked for completeness, was entered and coded using Epi Info version 5.0 and analyzed using Statistical Package for Social Sciences (SPSS) version 28. Continuous variables were presented as mean (SD) for normally distributed variables or median (interquartile range) for non-normally distributed variables. Categorical variables were reported as percentages and frequency tables. The Kaplan– Meier method estimated and graphed survival probabilities over time, and a log-rank hypothesis test to compare survival functions of explanatory variables was utilized. Significant predictor variables were identified by fitting a semiparametric Cox’s proportional hazard model using a method of forward stepwise and statistical significance variables were declared based on a p-value less than 0.05.

### 2.11 Ethical considerations

Proposal approval was obtained before the beginning of data collection from the Departmental Research and Promotion Committee (DRPC) of the Department of Internal Medicine, CHS. Permissions for data collection were obtained from the hospital administrative body. Confidentiality and anonymity of the client’s information were ensured throughout the execution of the study by taking only the required information without using the name of the client. Identification numbers were used rather than names to identify patients.

## 3. Result

### 1. Socio-demographic characteristics and lifestyle factors

Throughout the designated study period at Asella Referral and Teaching Hospital, 262 patients were admitted with the diagnosis of acute heart failure. Data was collected from 231 patients following the exclusion of 12 individuals based on predetermined criteria. Among those excluded, four patients exhibited stage 4 RVI, while three presented with advanced malignancies, including two cases of stage 4 cervical cancer and one case of stage 4 lung cancer. An additional five patients were discharged within 24 hours of admission, and records for 19 cases were inaccessible, having been misplaced from the medical record repository.

Within the group of 231 admitted patients with acute heart failure, more than half of the admitted patients were females, 121 (52.4%), and the majority, 167 (72.4%), were from rural residences. A significant portion, 142 (61.5%), of patients had active health insurance coverage. Twenty-seven individuals (11.6%) reported a history of cigarette smoking, and among them, 40.7% were identified as active smokers upon admission. For those classified as former smokers, the average duration since discontinuation was 2.9 years. Additionally, 22 participants (9.5%) had a history of alcohol intake, and a considerable portion of these individuals (81%) reported heavy alcohol consumption.

### 2. Clinical characteristics of patients at admission

Among the 231 patients admitted with acute heart failure during the study interval, more than half, 143 (61.9%), had worsening of preexisting chronic heart failure, and 88 (38.1%) had de novo heart failure at admission. Most patients, 187 (81.0%), presented with NYHA class IV, and 44 (19.0%) with NYHA class III. Upon admission, 224 (97%) were classified as having stage C heart failure, while 7 (3%) were categorized as stage D.

The hemodynamic profiles were as follows: 149 cases (64.5%) were warm and wet, 64 (27.7%) warm and dry, 13 (5.6%) cold and wet, and 5 (2.2%) cold and dry. The syndromic classification showed that 148 (64.1%) had acute decompensated heart failure (ADHF), 36 (15.6%) right-sided heart failure (RHF), 28 (12.1%) pulmonary edema (PE), and 19 (8.2%) cardiogenic shock (CS) (Table 2).

Importantly, echocardiographic assessment revealed that 117 patients (50.6%) had preserved ejection fraction (HFpEF), 87 (37.7%) had reduced ejection fraction (HFrEF), and 27 (11.7%) had mildly reduced ejection fraction (HFmrEF). This EF-based classification provides valuable insight into the disease profile and potential treatment implications of the cohort. (Table 6).

The mean systolic blood pressure (SBP) was 111 ± 27 mmHg, with 43 patients (18.61%) having a blood pressure below 90 mmHg. According to the ACC/AHA 2017 guidelines on hypertension, 128 patients (55.41%) out of the total 231 presented with a normal SBP upon admission. Additionally, 102 patients (44.2%) had an admission pulse rate exceeding 100 beats per minute. Thirty-eight individuals (16.45%) had a diastolic blood pressure below 60 mmHg, whereas the majority of patients (63.63%) presented with a normal diastolic blood pressure. Urine output data were documented for 201 patients, revealing a mean ± (SD) value of 2.1 ± 1.03 (Table 3).

More than half of these, 146 (63.2%), present with orthopnea and paroxysmal nocturnal dyspnea in 142 cases (61.47%), establishing these symptoms as the most prevalent Framingham major criteria in the study (Table 4).

Serum electrolyte levels were evaluated in 170 patients. The mean serum sodium level was 133.24 ± 8.8, wherein 97 individuals (42%) presented with hyponatremia, while 7 patients (3%) exhibited hypernatremia (>135 mEq/L). Similarly, serum potassium levels were determined for 170 patients, yielding a mean ± (SD) value of 3.7 ± 0.95. Among these, 78 individuals (33.8%) presented with hypokalemia, and 10 cases (4%) evidenced hyperkalemia (>5.5 mEq/L).

All 231 admitted patients had hemoglobin (Hgb) levels assessed, of whom 113 (48.9%) had anemia. Among these, 88 (77.9%) had moderate anemia, and 14 (12%) had severe anemia. Furthermore, among the 231 patients, serum creatinine levels were determined for 199 individuals, wherein 87 patients (43.7%) were diagnosed with acute kidney injury (AKI), with 17 cases (8.5%) developing AKI as a complication of treatment.

Chest X-ray findings were documented for 185 patients (80.09%). The most prevalent X-ray finding was pulmonary edema, observed in 70 cases (37.8%), followed by pneumonia in 45 instances (24.3%) and pleural effusion in 41 occurrences (22.1%).

All 231 patients underwent echocardiography, revealing ischemic heart disease in 84 cases (36.36%), chronic rheumatic heart disease in 66 patients (28.57%), and cor pulmonale in 44 instances (19.05%) as the three most prevalent findings.

Out of the 231 patients, only 149 individuals had an electrocardiogram. Atrial fibrillation was identified in 61 cases (40.9%), while 28 patients (18.8%) exhibited normal readings, and sinus tachycardia was noted in 20 individuals (13.4%).

Among the underlying cardiac conditions identified, ischemic heart disease was the most prevalent (84 cases) followed by chronic rheumatic heart disease (66 cases) and cor pulmonale (44 cases).

The three common precipitant factors that lead to hospitalization are pneumonia, atrial fibrillation, and noncompliance with medications, accounting for 84 cases (36.36%), 66 cases (28.57%), and 44 cases (19.05%), respectively.

Hypertension was the most common comorbidity identified, present in 54 (41.2%) cases, followed by diabetes in 32 (24.4%) cases and dyslipidemia in 25 (19%) cases.

As illustrated in figure 2, 64 patients (27.7%) developed treatment-related adverse drug effects during hospitalization, including hypokalemia (53.1%), acute kidney injury (AKI) (26.6%), and hyperkalemia (9.3%).

**Figure 1.**
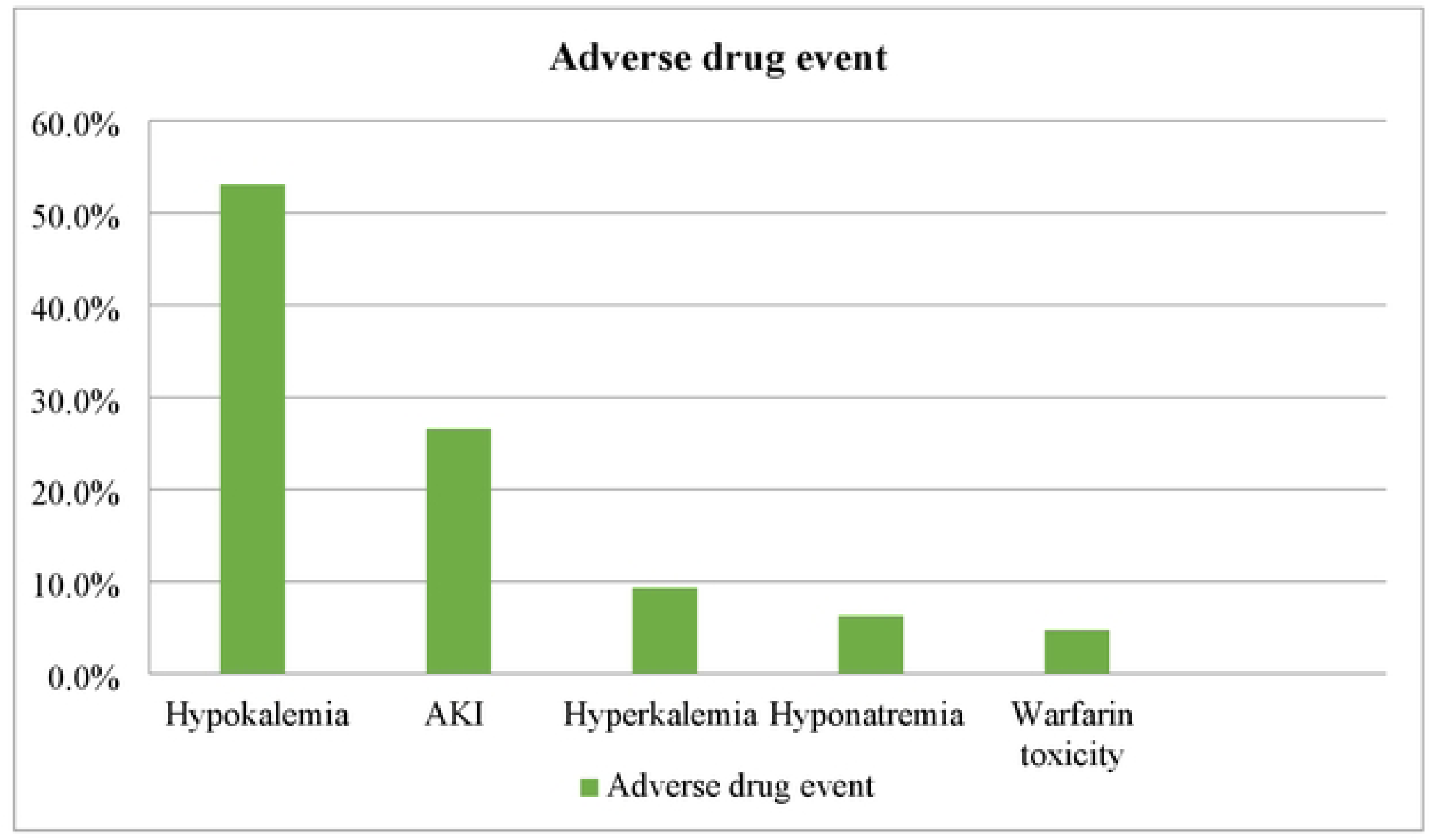
Adverse drug events among patients admitted with acute heart failure, ARTH, Ethiopia, 2024.

**Figure 2:**
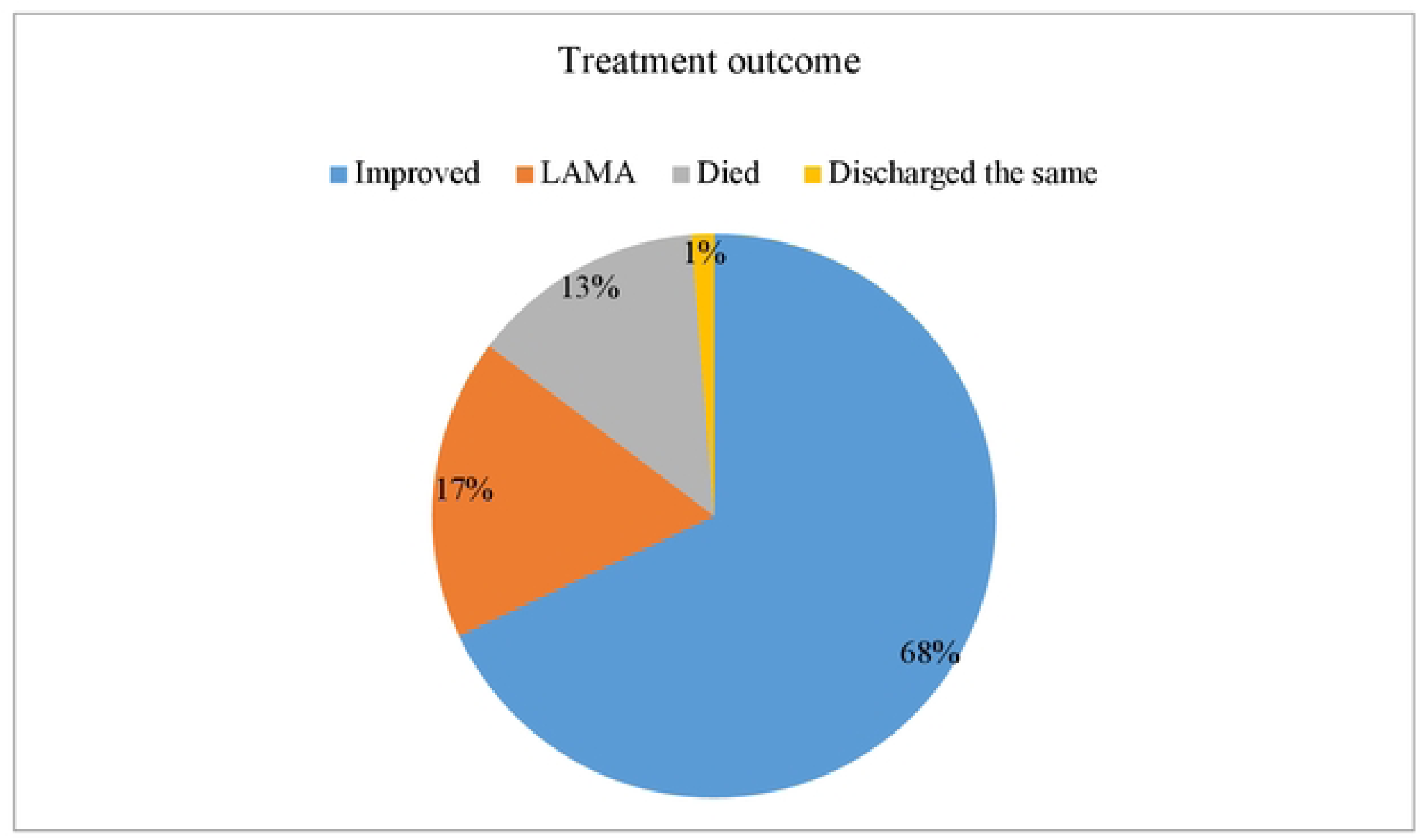
Treatment outcome of patients admitted with acute heart failure, **ARTH,** Ethiopia, 2024

### 3. Clinical condition at discharge and treatment outcome

At discharge, the majority of patients showed improvement in functional and hemodynamic status. Systolic blood pressure normalized in 137 patients (86.25%), and pulse rate was within normal limits in 127 patients (79.34%). There was also a marked shift in functional classification, with only two patients (1.25%) remaining in NYHA Class IV and 25 patients (15.63%) in Class III. The rest improved to Class II (48.13%) and Class I (35%). Additionally, most patients had no residual peripheral edema.

Regarding overall treatment outcome, 157 patients (68.0%) were discharged with clinical improvement, while 40 patients (17.3%) left against medical advice (LAMA), and 31 patients (13.4%) died during hospitalization (Figure 3).

**Figure 3:**
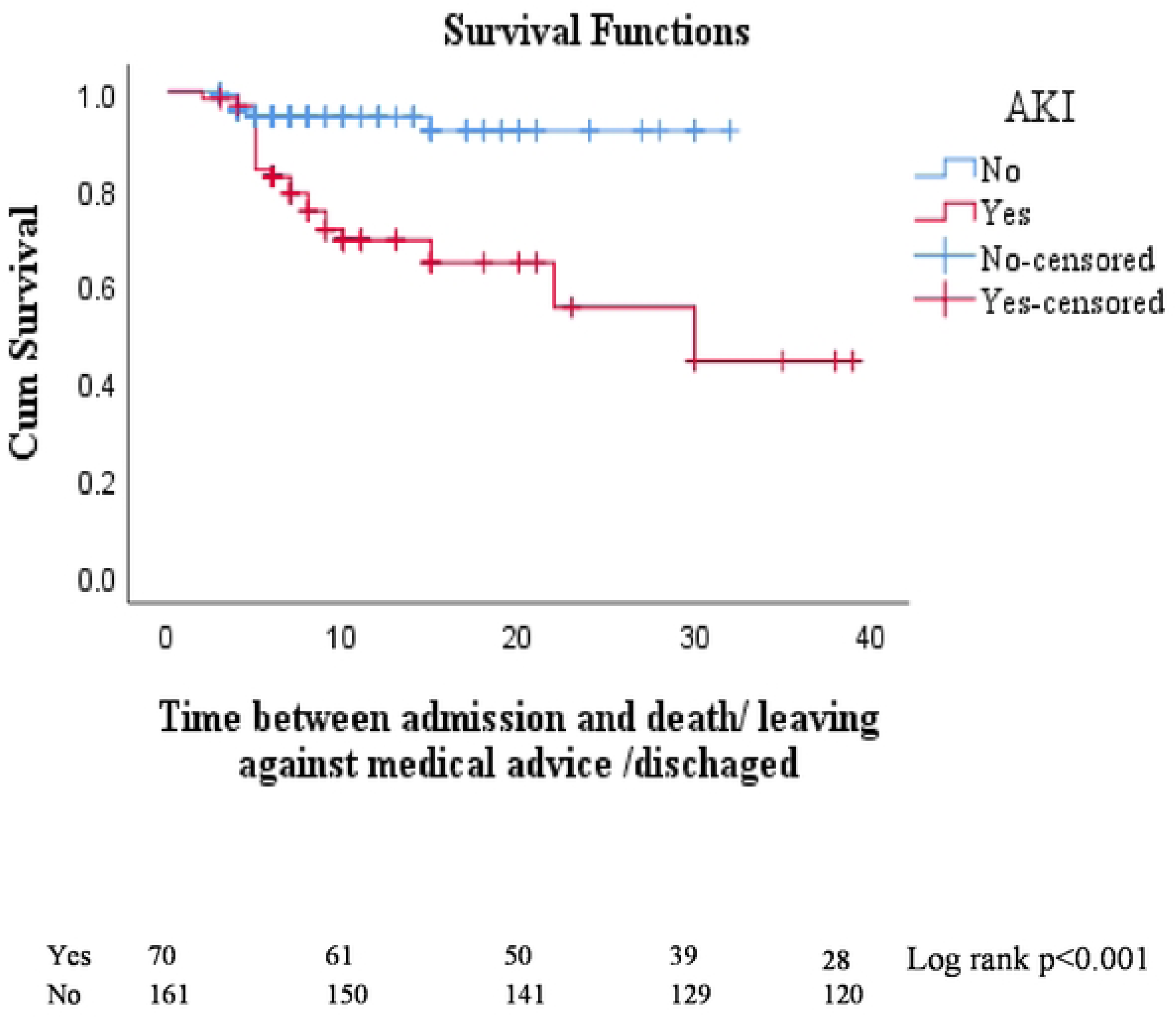
Kaplan-Meier of Acute kidney injury among patients admitted with acute heart failure, ARTH, Ethiopia, 2024.

**Figure 4:**
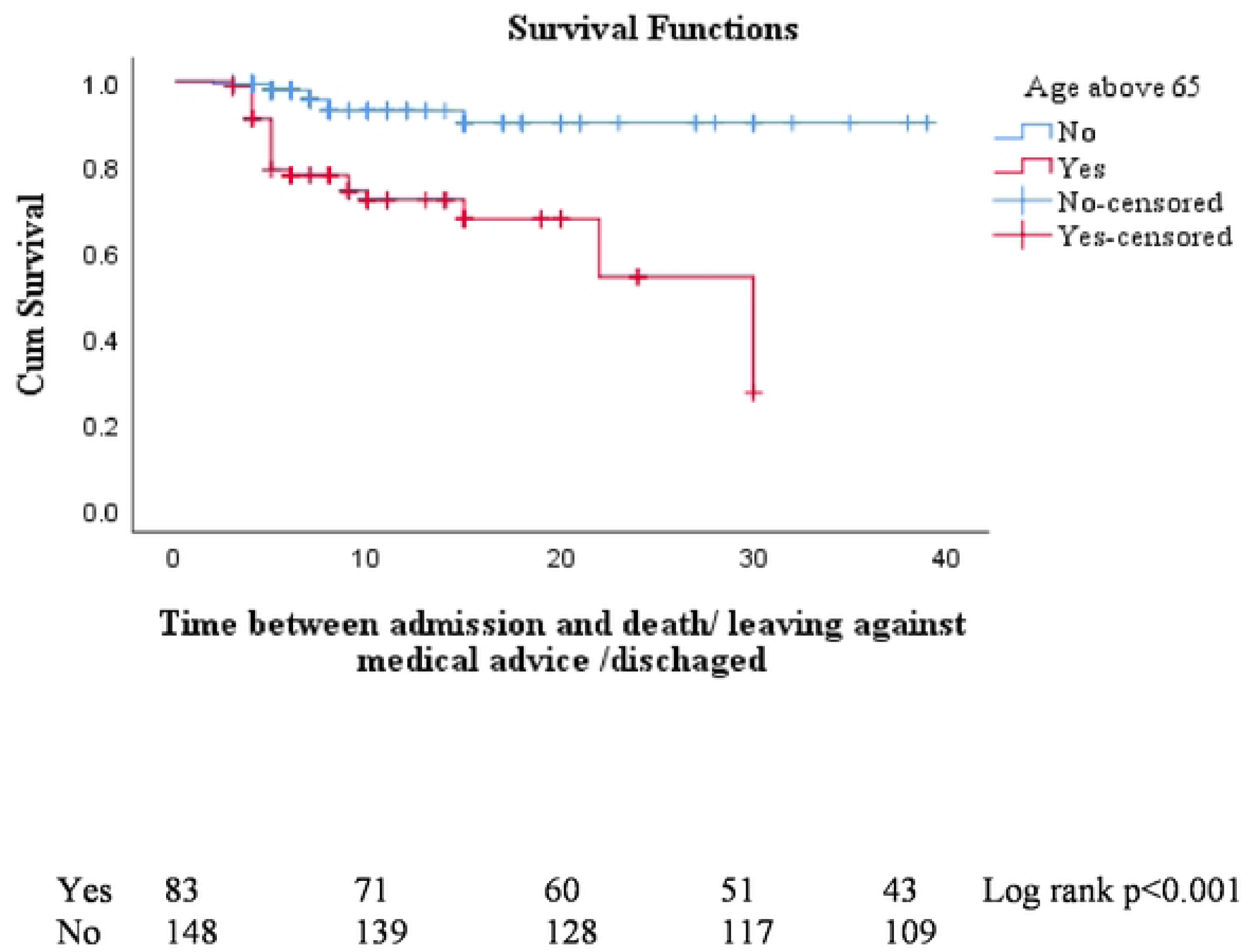
Kaplan-Meier of age among patients admitted with acute heart failure, ARTH, Ethiopia, 2024.

**Figure 5.**
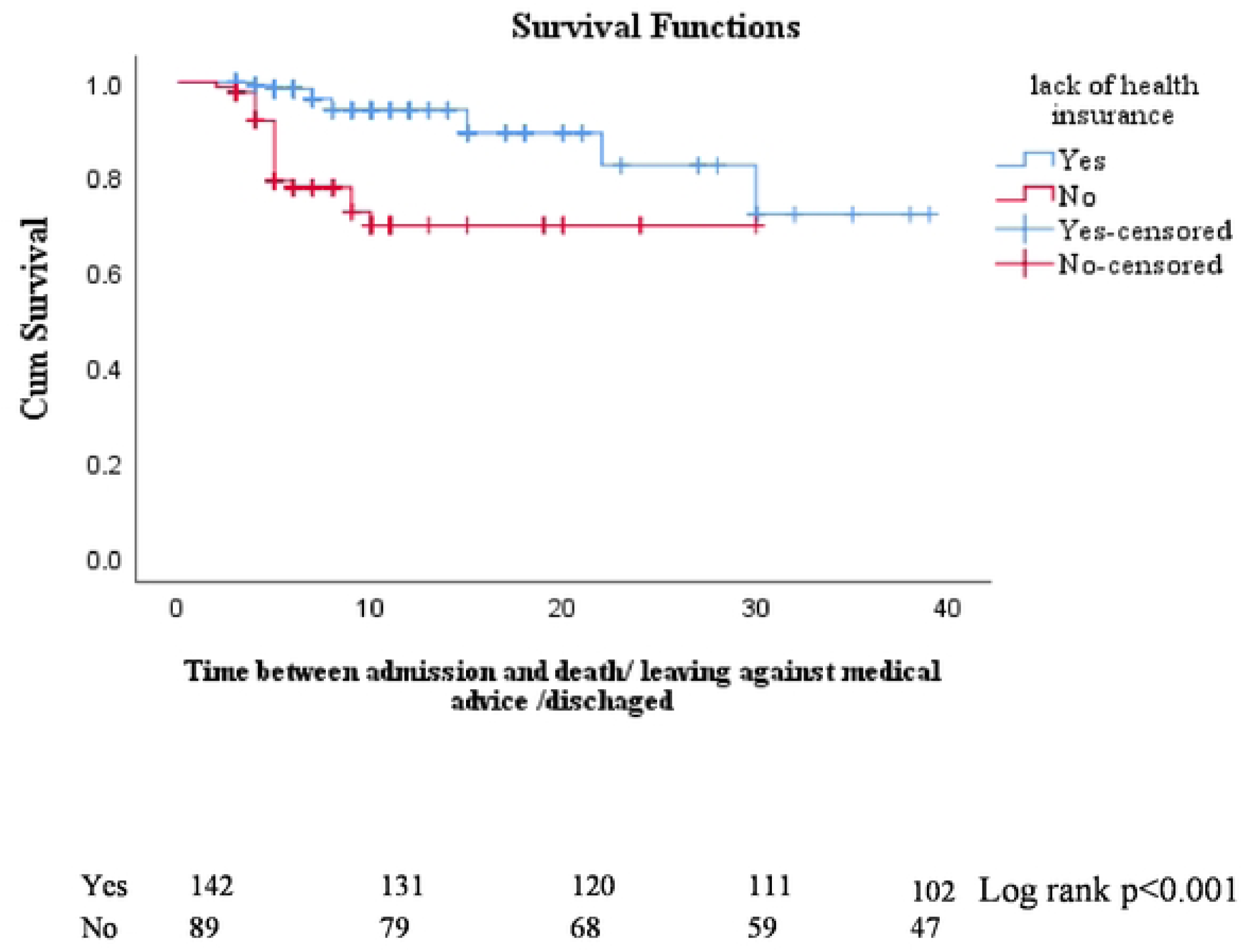
Kaplan-Meier of health insurance among patients admitted with acute heart failure, ARTH, Ethiopia, 2024.

**Figure 6:**
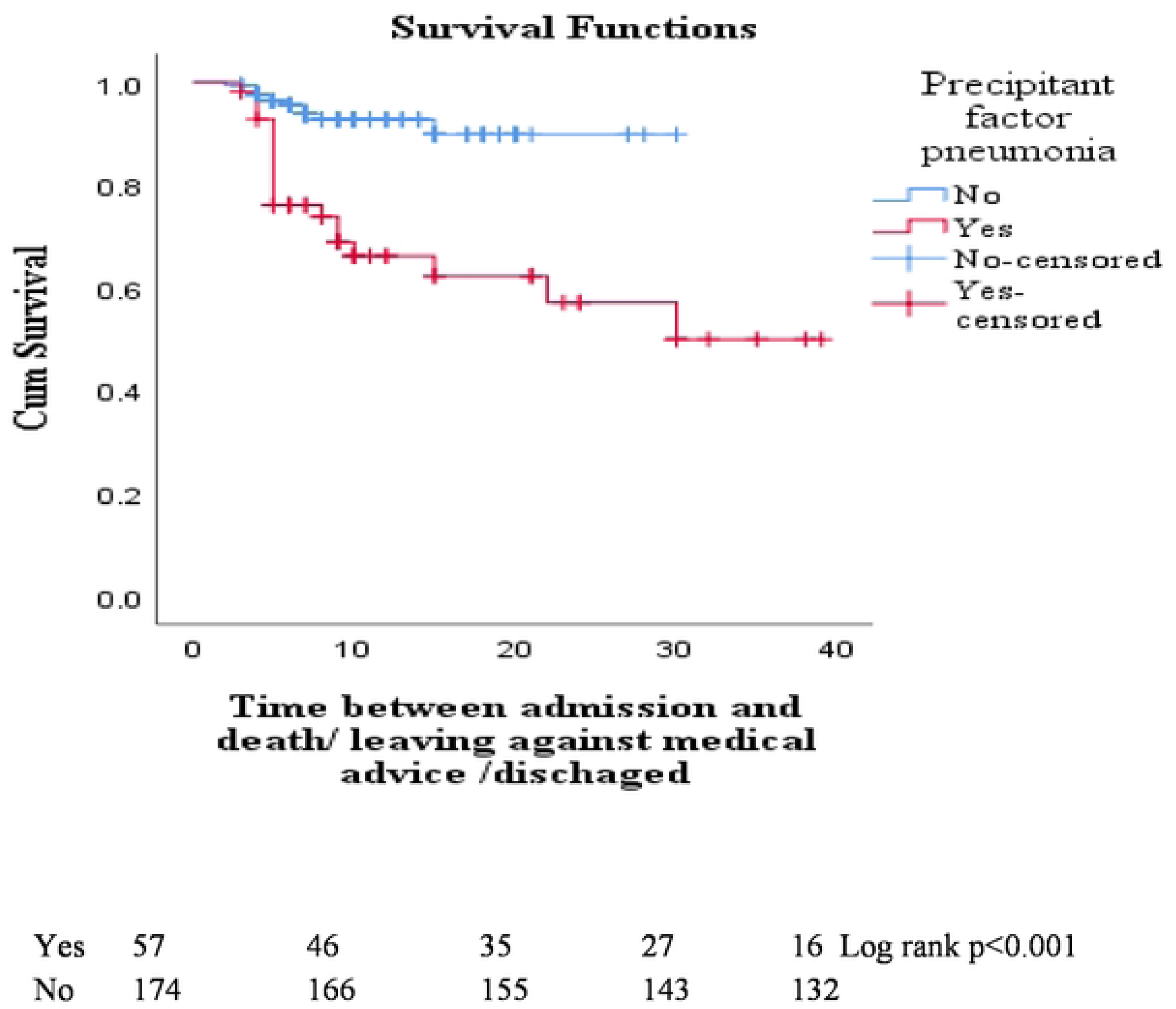
Kaplan-Meier of precipitant factor pneumonia among patients admitted with acute heart failure, ARTH, Ethiopia, 2024.

Notably, 27 patients (11.7%) were discharged in NYHA Class III or IV, indicating a persistent symptomatic burden and possibly unresolved congestion at the time of discharge. This may reflect advanced stage disease, limitations in inpatient optimization, or a need for closer post-discharge follow-up and palliative consideration in those with refractory symptoms, particularly the two individuals in Class IV.

### 4. Predictors of in hospital outcome

The Kaplan–Meier analysis, depicted in figures 3 to 7, demonstrates a clear difference of survival distributions in health insurance, age, acute kidney injury (AKI), and pneumonia. The log-rank test further supports these findings, revealing a highly significant p-value of <0.001.

The proportional hazard assumption was checked using the log-minus-log curve, and it does not violate the proportional assumption. The survival analysis has yielded insightful observations, emphasizing the impact of various factors on in-hospital outcomes. Pneumonia is identified as a substantial contributor, with a hazard ratio of 3.07 (95% CI: 1.40, 6.74, p = 0.005), indicating a significantly heightened risk. Similarly, acute kidney injury demonstrates a hazard ratio of 2.98 (95% CI: 1.31, 6.75, p = 0.009). Lack of health insurance also emerges as a noteworthy predictor, with a hazard ratio of 3.46 (95% CI: 1.51, 7.95, p = 0.003). Additionally, age, specifically age greater than 65, is identified as a pertinent factor, revealing a hazard ratio of 2.81 (95% CI: 1.18, 6.72, p = 0.020).

## 4. Discussion

Over half of the patients with acute heart failure (AHF) in this study were female, constituting 54.4% of the cohort. This female predominance is comparable to findings in other studies, such as the Acute Decompensated Heart Failure Registry (ADHERE) (52.0%) and the Organized Program to Initiate Lifesaving Treatment in Hospitalized Patients with Heart Failure (OPTIMIZE-HF) (52.0%). Notably, similar gender distribution patterns have been observed in studies from Tikur Anbessa Specialized (54.4%) and Jimma (53.3%) (26,27,49).

Regarding age, this study found that acute heart failure (AHF) predominantly affects a relatively young population, with a median age of 56 years. This is consistent with the Sub-Saharan Africa Survey of Heart Failure (THESUS–HF), which reported a median age of 55 years (6). In contrast, AHF in developed countries typically affects older individuals, with mean ages around 72 years and median ages between 66–70 years (26–28).

This age disparity can be attributed, in part, to differences in underlying etiologies. In our setting, chronic rheumatic heart disease (RHD)—a sequela of untreated streptococcal infections—is still prevalent and accounts for a significant proportion (28.6%) of AHF cases. RHD often leads to early-onset valvular dysfunction, resulting in symptomatic heart failure at a younger age. In contrast, heart failure in high-income countries is primarily driven by ischemic heart disease and lifestyle-related risk factors that manifest later in life. Furthermore, limited access to early surgical valve interventions and preventive care in low-resource settings contributes to progressive, irreversible cardiac damage in younger patients. These factors together help explain both the younger median age of AHF presentation and the persistence of advanced functional impairment at discharge in our cohort.

The majority of patients presented with worsening preexisting chronic heart failure (61.9%), with a notable proportion having de novo heart failure (38.1%). NYHA class IV was predominant at admission (81.0%), highlighting the severity of cases. A comparative study from Tikur Anbessa Specialized Hospital reported NYHA IV, III, and II at rates of 69.2%, 24.3%, and 6.5%, respectively (16). This disparity may be attributed to the higher representation of patients from rural areas in this study (72.4%), potentially contributing to delays in presentation with more severe or advanced decompensation.

The hemodynamic profiles indicated that a considerable number of patients presented with a warm and wet profile (64.5%), followed by warm and dry (27.7%). A study from Tikur Anbessa Specialized Hospital reported a similar pattern, with warm wet being the most common, followed by warm dry (16). The BREATHE registry also indicated warm and wet as the predominant hemodynamic profile (39).

The study showed that the most prevalent symptoms of Framingham major criteria were Orthopnea, paroxysmal nocturnal dyspnea, and rales. These findings are consistent with observations from the ALARM-HF registry, where orthopnea and rales emerged as the most common presentation (29).

In the present study, the principal precipitating factors identified included pneumonia, atrial fibrillation, and drug discontinuation. Similar patterns were observed in the BREATHE registry, where infection and poor medication adherence were prominent factors. These findings are also in concordance with the observations reported in the OPTIMIZE-HF registry, which highlighted pneumonia, arrhythmia, and acute coronary syndrome as the predominant precipitating factors (27,39). However, it is noteworthy that acute coronary syndrome (ACS) was less frequently identified as a precipitating factor in our study.

The findings suggest that implementing pneumonia vaccination programs, focusing on educating and counseling patients about cardiac drug compliance, along with optimizing the management of atrial fibrillation, might contribute to a decrease in admissions for acute heart failure.

In the majority of registries conducted in developed countries, heart failure with reduced ejection fraction (EF) is typically identified as the most type (27–29,40). However, in this study, heart failure with preserved ejection fraction is the predominant type, where despite a shift toward non-communicable diseases and ischemic heart disease becoming more prevalent, valvular and other infection-related conditions, such as pericarditis,. Continue to be predominant. This trend is evident in our study, where chronic rheumatic heart disease (CRHD) ranks as the second most common cause of heart failure.

In terms of underlying etiology, this study identified ischemic heart disease (IHD) as the most common cause of acute heart failure (36.36%), followed by chronic rheumatic heart disease (CRHD) and cor pulmonale. These findings are consistent with those reported in the OPTIMIZE-HF and BREATHE registries, as well as studies conducted in Djibouti and Jimma (21,27,39,49). In contrast, the THESUS–HF registry, conducted a decade ago, reported a predominance of non-ischemic etiologies, with hypertension and rheumatic heart disease being the leading causes (6). The difference between the two datasets likely reflects an ongoing epidemiological transition in sub-Saharan Africa. As highlighted in a recent study on the epidemiology of cardiovascular diseases in Morocco, there is a growing burden of cardiovascular risk factors such as diabetes, hypertension, dyslipidemia, and smoking, which may contribute to the increasing prevalence of ischemic heart disease in recent years (11).

Hypertension, diabetes mellitus, and dyslipidemia emerged as the three most common comorbidities identified in this research. A study conducted in Jimma reported similars (49). Likewise, in the KorAHF registry, major comorbidities included hypertension, diabetes, cerebrovascular illness, chronic renal failure, and chronicic lung disease (40). The impact of comorbidities on underlying cardiac disease was evident in this study, as approximately 44% of patients with ischemic heart disease had one or more comorbidities, whereas only 21% of patients with right-sided heart failure exhibited comorbidities. This underscores the significant influence of comorbidities on underlying cardiac conditions.

The in-hospital mortality rate was 13.4%, with higher mortality observed in patients with worsening of preexisting heart failure (61%) compared to those with de novo heart failure (39%). Overall, the in-hospital mortality in this study was lower than the mortality rate reported in the Jimma study (20.1%) and a prospective study conducted in Uganda (18.3%) (22,49). This difference could be attributed to improvements in health insurance coverage, as suggested by INTER-CHF, where lower healthcare insurance rates were identified as contributing factors for increased in-hospital mortality (8). Additionally, lack of health insurance coverage was identified as a predictor for poor in-hospital outcomes in this study. However, the in-hospital mortality rate among patients admitted to Assella Referral and Teaching Hospital remains relatively high compared to other registries such as KorAHF, OPTIMIZE-HF, and the ESC Heart Failure Long-Term Registry, where rates ranged from 4– 6.7% (27,28,40). As noted in a meta-analysis on heart failure care in low-and middle-income countries, this could be attributed to additional factors contributing to poor outcomes, including health-care infrastructure, quality of care, and variations in health-care access that may have contributed to higher mortality in our study (42).

The independent predictors of in-hospital mortality identified in this study included pneumonia, acute kidney injury, lack of active health insurance coverage, and age above 65. In alignment with these findings, the ADHERE registry reported that measurements of blood urea nitrogen (BUN), followed by systolic blood pressure (SBP) and serum creatinine at admission, were highly predictive of in-hospital mortality in patients with acute decompensated heart failure (ADHF) (26). Similarly, the OPTIMIZE-HF registry highlighted advanced age as a predictor of in-hospital outcomes (27).These consistent observations underscore the importance of these factors in assessing and predicting mortality risk in acute heart failure patients across different studies and registries.

## 5. Limitations of the study

The present study has several limitations that should be noted. Measurements of biomarkers such as B-type natriuretic peptide (BNP) and N-terminal pro-B-type natriuretic peptide (NT-pro-BNP) were not available, and these biomarkers could have provided important insights into predicting the outcome of acute heart failure (AHF).

Additionally, ECG and measurements of laboratory values, including cardiac troponin, renal function tests, and serum electrolytes, were not consistently available for all patients, and the absence of these data could affect the analysis and interpretation of outcomes. Moreover, it is important to acknowledge that the study is retrospective in nature, which may introduce inherent limitations.

## 6. Conclusion and Recommendation

### Conclusion

In summary, the study revealed a predominant representation of relatively young female patients experiencing acute decompensated heart failure (ADHF). Unfortunately, the in-hospital mortality rate for acute heart failure proved to be high in this study. The identified independent predictors of in-hospital mortality included pneumonia, acute kidney injury, lack of active health insurance coverage, low admission systolic blood pressures, and age above Ischemic heart disease emerged as the most common underlying cardiac disease, underscoring the significance of cardiovascular health in this population. The principal precipitating factors leading to hospitalization were pneumonia, atrial fibrillation, and drug discontinuation.

These findings underscore the complex interplay of demographic, clinical, and socioeconomic factors contributing to the outcomes of acute heart failure patients in our setting. The identification of specific predictors offers valuable insights for risk stratification and potential areas for targeted interventions.

### Recommendation

- Enhance Availability of Biomarkers: This study did not include measurement of key cardiac biomarkers such as B-type natriuretic peptide (BNP) or N-terminal pro-B-type natriuretic peptide (NT-proBNP) due to resource limitations. However, based on current international heart failure guidelines, improving the availability and accessibility of these biomarkers—especially in point-of-care settings—could significantly aid in the early diagnosis, risk assessment, and monitoring of acute heart failure (AHF) in similar low-resource environments.
- Implement Pneumococcal vaccination programs in rural clinics
- Improve consistency and availability of essential investigations such as electrocardiograms (ECGs) and laboratory values like cardiac troponin, renal function tests, and serum electrolytes for all patients to improve quality of care.
- Investigate targeted interventions to address independent predictors of in-hospital mortality, such as health insurance coverage.
- Tailored Interventions for Precipitating Factors: Design interventions addressing principal precipitating factors—pneumonia, atrial fibrillation, and drug discontinuation— through strategies for early detection, management, and patient education.

## Data Availability

All relevant data are within the manuscript and its Supporting Information files.

## 7. Lists of Abbreviations/ Acronyms

ACC/AHA: American College of Cardiology / American Heart Association
ACE: Angiotensin Converting Enzyme
ACS: Acute Coronary Syndrome
ADHERE: Acute Decompensated Heart Failure National Registry
ADHF: Acute Decompensated Heart Failure
ADR/SE: Adverse Drug Event / Side Effect
AF: Atrial Fibrillation
AHF: Acute Heart Failure
ALARM-HF: Acute Heart Failure Global Registry of Standard Treatment
ATRH: Asella teaching and referral hospital
BNP: B-type Natriuretic Peptide
BUN: Blood Urea Nitrogen
CAD: Coronary Artery Disease
CHD: Congenital Heart Disease
CKD: Chronic Kidney Disease
COPD: Chronic Obstructive Pulmonary Disease
CHS: College of Health Sciences
CS: Cardiogenic shock
DCM: Dilated Cardiomyopathy
DM: Diabetes Mellitus
ED: Emergency Department
EF: Ejection Fraction
EHFS: II EuroHeart Failure Survey II
ESC: European Society of Cardiology
ESC-HF: pilot EURObservational Research Program the Heart Failure Pilot Survey
EPHI: Ethiopian Public Health Institute
HDU: High dependency unit
HF: Heart Failure
HHD: Hypertensive Heart Disease
HIV/AIDS: Human Immunodeficiency Virus / Acquired Immune Deficiency Syndrome
ICU: Intensive Care Unit
IHD: Ischemic Heart Disease
INTER-CHF: International Congestive Heart Failure III
IQR: Interquartile range
KorAHF: Korean Acute Heart Failure Registry
LAMA: Left against medical advice
LVEF: Left Ventricular Ejection Fraction
LVH: Left Ventricular Hypertrophy
MDRD: Modification of Diet on Renal Disease
MI: Myocardial Infarction
MR: Mitral Regurgitation
NTproBNP: N-terminal pro-B-type Natriuretic Peptide
NYHA: New York Heart Association
OPTIMIZE-HF: Organized Program to Initiate Lifesaving Treatment in Hospitalized Patients with Heart Failure
PE: Pulmonary edema
RHD: Rheumatic Heart Disease
RHE: Right side heart failure
SBP: Systolic Blood Pressure
SSA: Sub-Sahara Africa
TASH: Tikur Anbessa Specialized Hospital
THESUS–HF: The Sub-Saharan Africa Survey of Heart Failure
TR: Tricuspid Regurgitation
VHD: Valvular Heart Disease

## 8. Acknowledgements

We are grateful to thank the study participants and their health personnel.

## 9. Authors’ contributions

Duresa Mieso Elemo: Conceptualization, Methodology, Investigation, Data curation, Formal analysis, Writing - original draft.

Wubshet Abraham Alemu: Supervision, Project administration, Methodology, Formal analysis, Writing - review & editing.

Tamiru Adugna Dadi: Conceptualization, Data curation, Validation, Formal analysis, Writing - review & editing.

Koricho Simie Tolla: Investigation, Resources, Writing - review & editing.

Mengesha Akale Tekle: Formal analysis, Visualization, Writing - review & editing.

Ayalneh Demissie Ashenafi: Conceptualization, Data curation, Validation, Formal analysis, Writing - review & editing.

All authors participated in study design, data interpretation, and critical revision of the manuscript. Each author approved the final version and agrees to be accountable for all aspects of the work

## Funding

The authors received no specific funding for this work.

## Availability of data and materials

All relevant data are within the manuscript and its Supporting Information files.

## Ethics approval and consent to participate

Ethical approval was obtained from the Institutional Review Board (IRB) of Arsi University, College of Health Sciences. Protocol No: **A/CHS/RC/68/2023**. Formal permission letter was obtained from hospital authorities.

## Consent for publication

Not applicable

## Data availability

The datasets used or analyzed during the current study are available from the corresponding author upon reasonable request.

## Competing interests

The authors declare no competing interests.

## Notes

### Competing Interest Statement

The authors have declared no competing interest.

### Funding Statement

The author(s) received no specific funding for this work.

### Author Declarations

Ethical approval was obtained from the Institutional Review Board (IRB) of Arsi University, College of Health Sciences. Reference number (A/U/H/S/C/120/8286/2024). A formal permission letter was obtained from hospital authorities.

